# Kinetics and seroprevalence of SARS-CoV-2 antibodies – a comparison of 3 different assays

**DOI:** 10.1101/2021.03.10.21253273

**Authors:** Elisabeth Kahre, Lukas Galow, Manja Unrath, Luise Haag, Judith Blankenburg, Alexander H. Dalpke, Christian Lück, Reinhard Berner, Jakob P. Armann

## Abstract

**Purpose:** Comparing seroprevalence and antibody kinetics in three different commercially available assays for SARS-CoV-2.

**Methods:** Serostatus of COVID-19 patients was analyzed 5 months and 10 months after their infection, using three different assays: Diasorin LIAISON®, Euroimmun®, Abbott Diagnostics® ARCHITECT.

**Results:** Seropositivity at baseline differed significantly depending on the assay (Diasorin 81%, Euroimmun 83%, Abbott 59%). At follow-up antibody levels detected in the Diasorin assay were stable, while there was a significant loss in seropositivity in the Euroimmun and Abbott assays.

**Conclusion:** There are significant differences in SARS-CoV-2 antibody kinetics based on the specific assay used.

**Trial registration number, date of registration:** DRKS00022549, 29.07.2020 “retrospectively registered”

## Introduction

Since the beginning of the SARS-CoV-2-pandemic [1] more than 112 Million cases and almost 2.5 Million [2] deaths have been reported worldwide. Serum antibody testing is becoming a critical tool, both in diagnosis of COVID-19 and in seroprevalence studies.

Most patients develop detectable antibodies within 14 days after their infection [3]. However, there is inconsistent evidence regarding the duration of antibody persistence with studies showing only short lived antibody responses [4] while others showing persistent serum levels [5].

While differences in patient characteristics and disease course might explain some contradictory findings, assay dependent differences might also play a role.

We analyzed SARS-CoV-2 seroprevalence and antibody kinetics over 9 months in 109 individuals using three different commercially available assays.

## Methods

### Study Design

Patients with PCR confirmed SARS-CoV-2 infection in Dresden (a city in Saxony/Germany with approximately 557.000 inhabitants) were invited via the local health department to participate in the AmbCoviDD19 study.

After informed consent was obtained, 5 mL of peripheral venous blood was collected from each individual to assess SARS-CoV-2 IgG antibodies at baseline and 9-12 months after their infection (follow-up). Additional information about age, comorbidities, regular medication, COVID-19-symptoms, disease course and test indication were obtained.

The AmbCoviDD19 study was approved by the Ethics Committee of the Technische Universität (TU) Dresden (BO-EK-137042020) and has been assigned clinical trial number DRKS00022549.

### Laboratory Analysis

We assessed SARS-CoV-2 IgG antibodies in all samples using three commercially available assays.

First, chemiluminescence immunoassay (CLIA) technology for the quantitative determination of anti-S1 and anti-S2 specific IgG antibodies to SARS-CoV-2 was used: Diasorin LIAISON^®^ SARS-CoV-2 S1/S2 IgG Assay. Antibody levels > 15.0 AU/ml were considered positive and levels between 12.0 and 15.0 AU/ml were considered equivocal.

Second, an ELISA detecting IgG against the S1 domain of the SARS-CoV-2 spike protein, Euroimmun^®^ Anti-SARS-CoV-2 ELISA, was used; a ratio < 0.8 was considered negative, 0.8– 1.1 equivocal, > 1.1 positive.

Third, chemiluminescent microparticle immunoassay (CMIA) intended for the qualitative detection of IgG antibodies to the nucleocapsid protein of SARS-CoV-2, Abbott Diagnostics^®^ ARCHITECT SARS-CoV-2 IgG, was used; an index (S/C) of < 1.4 was considered negative, >/= 1.4 was considered positive.

### Statistical Analysis

Analyses were performed using IBM SPSS 25.0 and Microsoft Excel 2010. Statistical comparisons between groups were performed using the Fishers’ exact test for categorical variables and T-test for means. Correlations were assessed using a Spearman’s Rank correlation coefficient (r). All tests were 2-sided, and *p* values⍰<⍰0.05 were considered statistically significant.

### Funding

This study was supported by a grant of the Federal State of Saxony, Germany.

## Results

Overall, 109 individuals with a positive PCR in respiratory samples between March and May 2020 were enrolled in this study. 57/109 (52%) were female, median age was 46 years, 2/109 (1.8%) were younger than 19 years, 8/109 (7.3%) required hospitalization and 92/109 (84%) reported COVID-19 related symptoms (see Table 1 for full patients’ characteristics).

**Table 1.** Patients’ characteristics

|  |  |  |
| --- | --- | --- |
| **Age (years)** | Mean (Range) | 46 (4-80) |
| **Gender** | Female | 57 (52%) |
| **Symptomatic SARS-CoV-2 infection** |  | 92 (84%) |
| **Hospitalization** |  | 8 (7%) |
| **Time interval from PCR to baseline (days/months)** | Median | 144/4.8 |
|  | IQR | 127-154/4.2-5.1 |
| **Time interval from PCR to follow-up (days/months)** | Median | 295/9.7 |
|  | IQR | 288-301/9.5-9.9 |

*IQR*, interquartile range

Median time between first serological testing and positive PCR was 144 days (4.8 months).

92/109 (84%) participants had detectable antibodies against SARS-CoV-2 in at least one assay; 88/109 (81%) were Diasorin positive while 90/109 (83%) Euroimmun positive and 64/109 (59%) Abbott positive.

88/109 (81%) participants had detectable antibodies against SARS-CoV-2 in at least two different assays at baseline. Of these, 1/88 (1%) had no detectable antibodies in the Diasorin, 25/88 (28%) in the Abbott whereas all of these participants had at least equivocal results in the Euroimmun assay. 62/109 (57%) individuals had detectable antibodies in all three assays at baseline (Table 2 a, Fig. 1).

**Table 2.** Seroprevalence at baseline and follow-up.

**a) Comparison of assays between baseline and follow-up**
|  | Baseline | Follow-up | *p* value |
| --- | --- | --- | --- |
| Diasorin LIAISON® | 88/109 (81%) | 91/109 (83%) | n.s. |
| Euroimmun® ELISA | 90/109 (83%) | 85/109 (78%) | n.s. |
| Abbott Architect® | 64/109 (59%) | 26/109 (24%) | 0.0001 |
| Seropositive in one assay | 92/109 (84%) | 101/109 (93%) | n.s. |
| Seropositive in two assays | 88/109 (81%) | 77/109 (71%) | n.s. |
| Seropositive in three assays | 62/109 (57%) | 24/109 (22%) | 0.0001 |

**b) Development of antibodies from baseline to follow-up**
|  | Baseline | Follow-up | *p* value |
| --- | --- | --- | --- |
| Diasorin | 88 | 86 | n.s. |
| Euroimmun | 90 | 73 | 0.0004 |
| Abbott | 64 | 19 | 0.0001 |

**Fig. 1.**
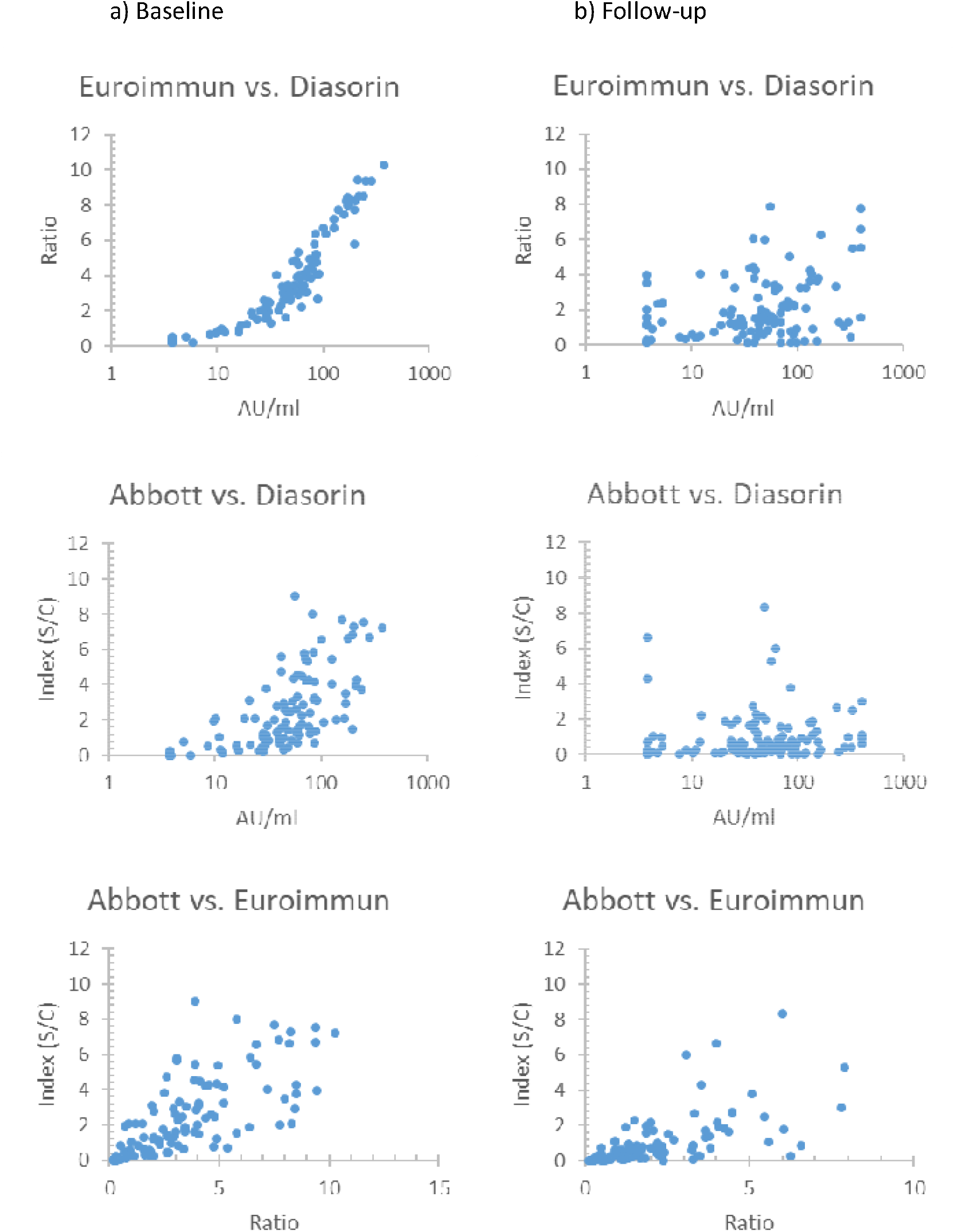
Relationship between levels of antibodies in different assays

Time between infection and serological testing (146 days vs. 141 days), age (44 vs. 46 years), female gender (51% vs. 57%) hospitalization rate (8/88 (9%) vs. 0/21 (0%)) and frequency of reported symptoms (75/88 (85%) vs. 17/21 (81%)) did not differ significantly between seropositive and seronegative individuals (Table 3).

**Table 3.** Factors not influencing seropositivity at baseline.

| **Factors** | ≥ 2 tests positive | ≥ 1 test negative | *p* value | ≥ 1 test positive | No test positive | *p* value |
| --- | --- | --- | --- | --- | --- | --- |
| **Female gender** | 45/88 (51%) | 12/21 (57%) | n.s. | 48/92 (52%) | 9/17 (53%) | n.s. |
| **Age** | 44 (32-58) | 46 (34-60) | n.s. | 43 (31-58) | 49 (40-60) | n.s. |
| **Time interval from PCR to baseline** | 146 (129-156) | 141 (97-149) | n.s. | 147 (129-156) | 138 (91-148) | n.s. |
| **Symptomatic** | 75/88 (85%) | 17/21 (81%) | n.s. | 77/92 (84%) | 15/17 (88%) | n.s. |
| **Hospitalization** | 8/88 (9%) | 0/21 (0%) | n.s. | 8/92 (9%) | 0/17 (0%) | n.s. |

Seropositivity, however, differed significantly depending on test indication. While 44/50 (88%) participants with a defined SARS-CoV-2 positive contact were seropositive, 18/19 (95%) were positive with a travel history, only 16/27 (59%) were seropositive when tested solely based on symptoms (Fig. 2).

**Fig. 2.**
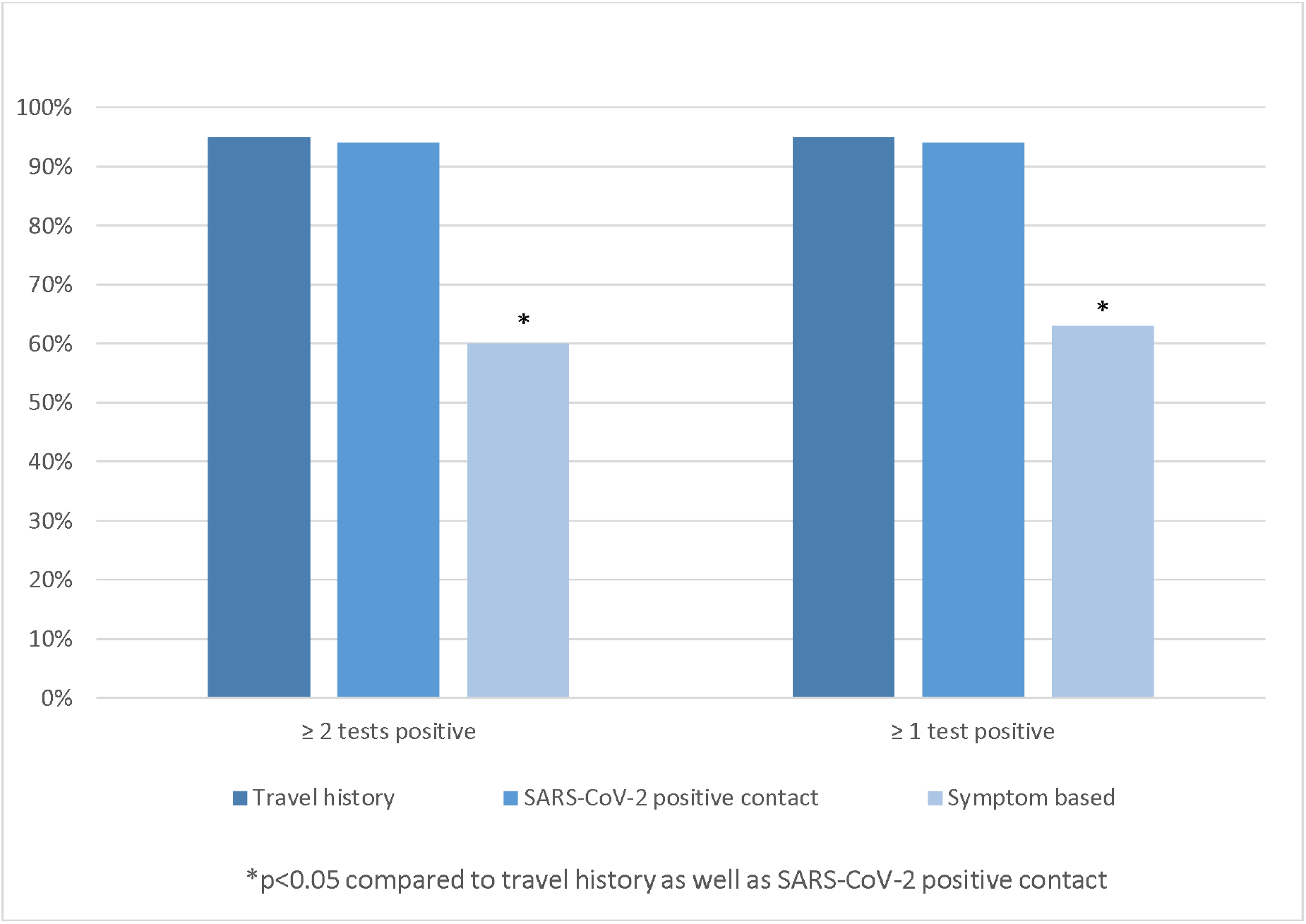
Factors influencing seropositivity; *p* value refer to comparison between travel history and symptom based test indication as well as between SARS-CoV-2 positive contact and symptom based test indication

Median time between second antibody assessment (follow-up) and infection was 295 days (9.8 months) (Table 1). 101/109 (93%) participants had detectable antibodies in at least one assay. 89/92 (97%) with detectable antibodies at baseline in at least one assay continued to have detectable antibodies in at least one assay at follow-up. 91/109 (83%) were Diasorin positive, 85/109 (78%) Euroimmun positive and 26/109 (24%) Abbott positive (Table 2 a).

However, while 86/88 (98%) of participants with initially detectable antibodies in the Diasorin assay continued to have detectable antibodies in this assay, numbers were significantly lower for the Euroimmun (73/90 (81%) *p* 0.0004) and the Abbott (19/64 (30%) p 0.0001) (Table 2 b). In addition mean antibody levels were stable in the Diasorin assays over time while they dropped significantly in the Euroimmun and Abbott assays (Fig. 3).

**Fig. 3.**
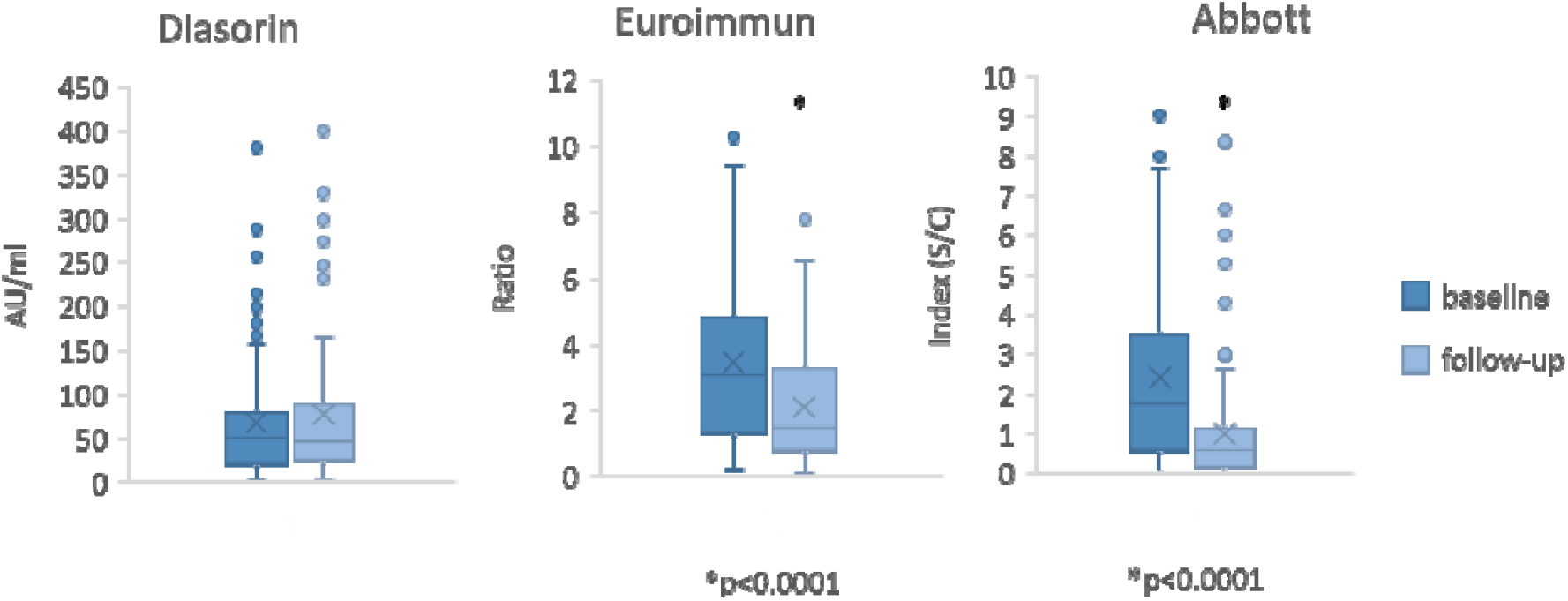
Mean antibody levels over time in different assays; *p* value refer to comparison between baseline and follow-up

At follow-up, 77/109 (71%) had detectable antibodies in at least two different assays (Table 2 a); 71/88 (81%) individuals with two positive serological tests at baseline continued to have detectable antibodies in two assays; 15/17 (88%) individuals no longer tested positive in two different assays continued to have detectable antibodies in the Diasorin, while none of these continued to have detectable antibodies in the Euroimmun or Abbott. 24/109 (22%) had detectable antibodies in all three assays at follow-up (Table 2 a, Fig. 1).

Five out of 21 (24%) individuals without detectable antibodies in the Diasorin assay at baseline had detectable antibodies at follow-up, 7/19 (37%) in the Euroimmun and 7/45 (16%) in the Abbott respectively. Of the 17 individuals without detectable antibodies in any assay at baseline, 12 (71%) had detectable antibodies in at least one assay at follow-up (3 Diasorin / 10 Euroimmun / 3 Abbott). Two individuals reported a positive SARS-CoV-2 PCR testing between baseline and follow-up. Both had initially no detectable antibodies in any assay, one had antibodies in all three assays at follow-up, one had detectable antibodies in the Diasorin and Euroimmun. Five individuals had no detectable antibodies in any assay at both times. Four out of five were tested by PCR based on symptoms alone, while 1/5 was tested due to travel history.

## Discussion

The overall seroconversion rate in our study is comparable to large population based seroprevalence studies [6, 7], however the differences in seroprevalence detected by different assays are striking. While at baseline the Diasorin and Euroimmun provide comparable results (Fig. 1), the seroprevalence in the Abbott is 20% lower compared to the other assays. At follow-up these differences become even more pronounced. While the Diasorin shows stable persistent antibody levels, seropositive rates detected with Euroimmun and Abbott decrease significantly over time, leading to a possible underestimation of seropositivity of up to 60%. These differences might at least partly explain contradicting findings in longitudinal antibody kinetic studies.

Given the intra-individual differences, patient characteristics seem unlikely to explain the discrepancies between the different assays. Differences in antibody persistence based on targeted epitopes might play a role comparing Diasorin and Euroimmun with Abbott. However, the significant difference between Diasorin and Euroimmun at follow-up - both detecting antibodies targeting the spike protein of SARS-CoV-2 – requires further investigation.

In addition, antibodies to the nucleocapsid protein of SARS-CoV-2 are thought to be useful in differentiating between seroconversion after infection and vaccination. Given the substantial loss of these antibodies over time in our sample, this approach might be less feasible than expected.

It is remarkable that 71% of individuals without any detectable SARS-CoV-2 antibodies 4-5 months after their infection had measurable levels later at 9-10 months. One possible explanation could be that their initial short lived antibody response was boostered during the study period through repeat exposure. Further immunological studies – including T-cell assays – are needed to investigate this further. The observation that the only PCR confirmed repeat infections occurred in individuals without a detectable antibody response at baseline is somewhat reassuring and might point to a correlation between antibody response and immunity.

The lack of significant associations between seropositivity and patient characteristics in our study compared to previous [5,7] might be explained by the relatively long time period between infection and baseline serological testing as well as the rather homogenous study population with mild clinical courses.

The significant differences in seropositivity based on test indication is of particular interest though and most likely due to an increased rate of false positive PCRs in populations with a lower pre-test-probability. While this association is well described in the literature [8] these results are an important reminder that even highly specific test as the SARS-CoV-2 RT-qPCR do have a measurable false positive rate [9]. This observation needs to be given special consideration when implementing population-based screening programs in asymptomatic individuals.

## Conclusion

There are significant differences in SARS-CoV-2 antibody kinetics based on the specific assay used. In our cohort the Diasorin LIAISON^®^ assay performed best regarding long term detection of seropositivity. There is a significant difference in seropositivity in PCR-positive individuals based on the indication for PCR-testing.

## Data Availability

We share data if reasonable requests are received. Requests should be directed to the corresponding author at

## Declarations

## Funding

This study was supported by a grant by the Federal State of Saxony.

## Conflicts of interest/Competing interests

Alexander Dalpke, Reinhard Berner and Jakob Armann report grants from Federal State of Saxony during the conduct of the study.

## Code availability

N/A

## Authors’ contributions

Jakob Armann, Lukas Galow, Elisabeth Kahre, Reinhard Berner and Alexander Dalpke designed the study and wrote the protocol. Jakob Armann, Lukas Galow, Manja Unrath, Judith Blankenburg, Elisabeth Kahre and Luise Haag collected samples. Alexander Dalpke and Christian Lück performed serological testing. Jakob Armann, Lukas Galow, Elisabeth Kahre and Reinhard Berner analyzed and verified the data. Elisabeth Kahre, Lukas Galow and Jakob Armann wrote the manuscript. Alexander Dalpke, Christian Lück and Reinhard Berner reviewed the manuscript.

## Notes

### Clinical Trial

DRKS00022549

### Author Declarations

The AmbCoviDD19 study was approved by the Ethics Committee of the Technische Universitaet (TU) Dresden (BO-EK-137042020)

## REFERENCES

1 Zhu N, Zhang D, Wang W, et al. A Novel Coronavirus from Patients with Pneumonia in China, 2019. N Engl J Med 2020;382(8):727–33. doi:10.1056/NEJMoa2001017 [published Online First: 24 January 2020].

2 European Centre for Disease Prevention and Control. COVID-19 situation update worldwide, as of week 7, updated 25 February 2021 2021. Available at: https://www.ecdc.europa.eu/en/geographical-distribution-2019-ncov-cases Accessed February 28, 2021.

3 Sun B, Feng Y, Mo X, et al. Kinetics of SARS-CoV-2 specific IgM and IgG responses in COVID-19 patients. Emerg Microbes Infect 2020;9(1):940–48.

4 Carsetti R, Zaffina S, Piano Mortari E, et al. Different Innate and Adaptive Immune Responses to SARS-CoV-2 Infection of Asymptomatic, Mild, and Severe Cases. Front Immunol 2020;11:610300. doi:10.3389/fimmu.2020.610300 [published Online First: 16 December 2020].

5 Zhang X, Lu S, Li H, et al. Viral and Antibody Kinetics of COVID-19 Patients with Different Disease Severities in Acute and Convalescent Phases: A 6-Month Follow-Up Study. Virol Sin 2020;35(6):820–29. doi:10.1007/s12250-020-00329-9 [published Online First: 22 December 2020].

6 Pollán M, Pérez-Gómez B, Pastor-Barriuso R, et al. Prevalence of SARS-CoV-2 in Spain (ENE-COVID): a nationwide, population-based seroepidemiological study. The Lancet 2020;396(10250):535–44.

7 Gudbjartsson DF, Norddahl GL, Melsted P, et al. Humoral Immune Response to SARS-CoV-2 in Iceland. N Engl J Med 2020;383(18):1724–34. doi:10.1056/NEJMoa2026116 [published Online First: 1 September 2020].

8 Jackson BR. The dangers of false-positive and false-negative test results: false-positive results as a function of pretest probability. Clin Lab Med 2008;28(2):305-19, vii.

9 Surkova E, Nikolayevskyy V, Drobniewski F. False-positive COVID-19 results: hidden problems and costs. The Lancet Respiratory Medicine 2020;8(12):1167–68.

